## Supplementary figures and images for "Identifying metabolic features of colorectal cancer liability using Mendelian randomization"

### Figure 2-figure supplement 1

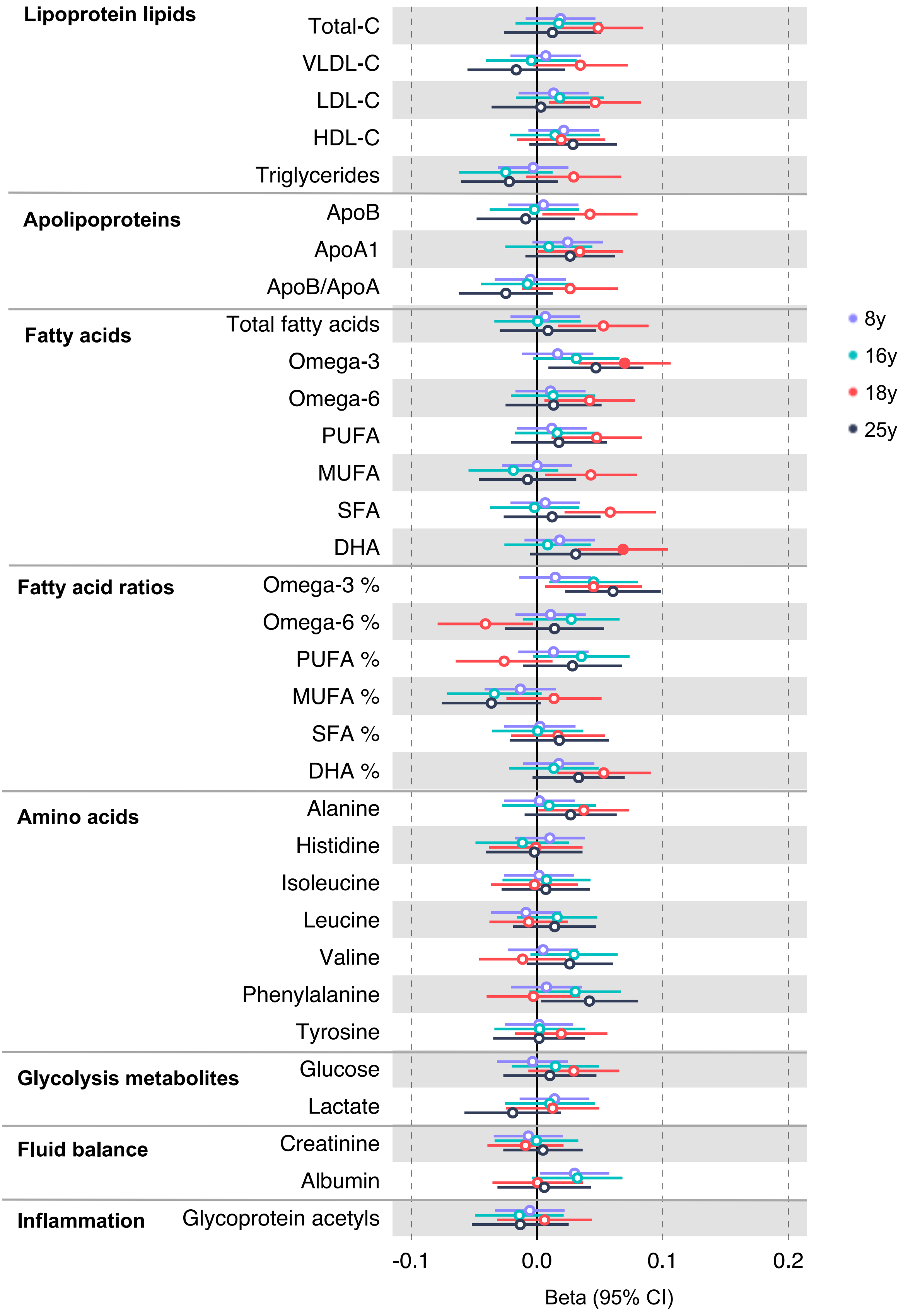

### Figure 2-figure supplement 2

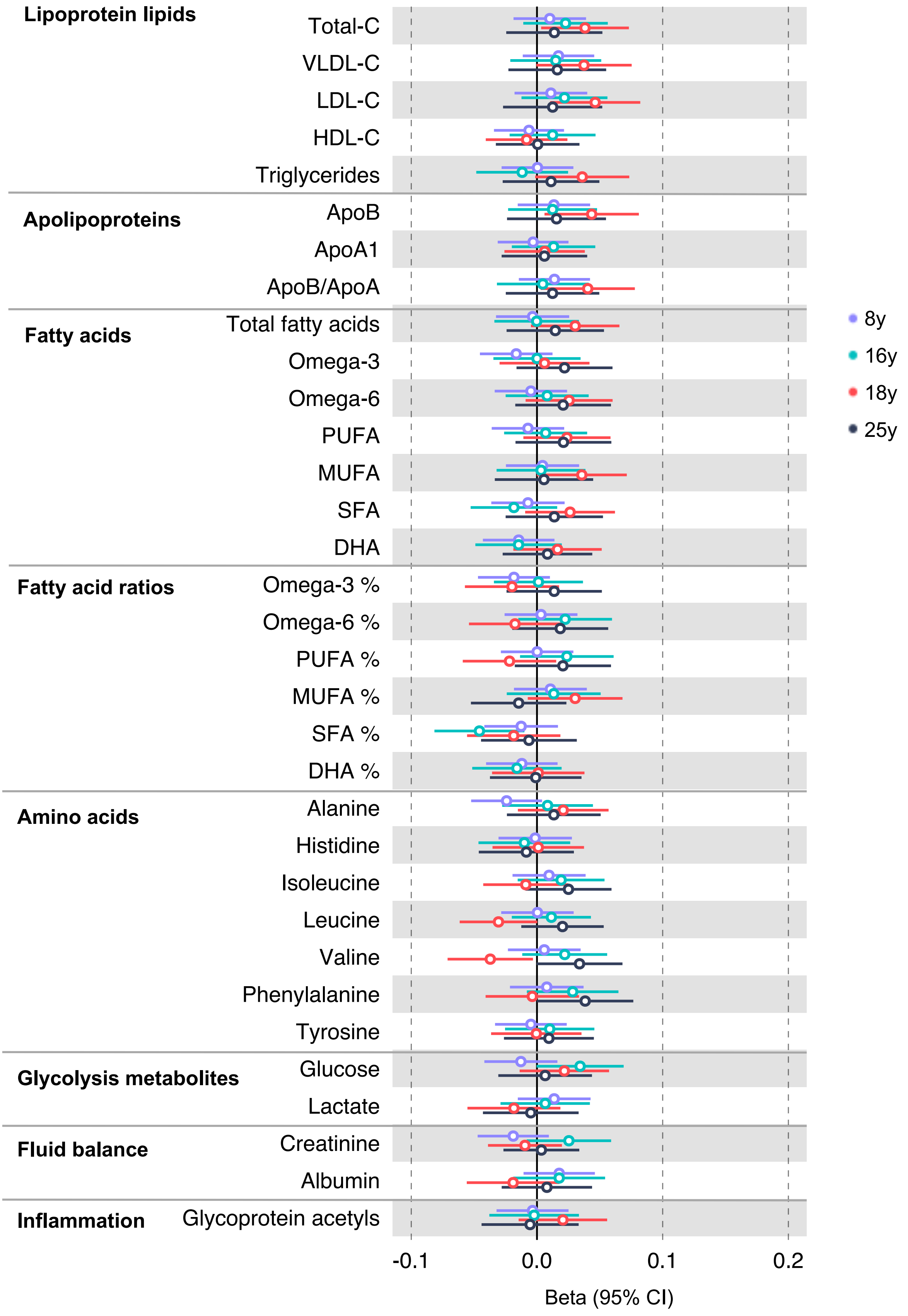

### Figure 2-figure supplement 3

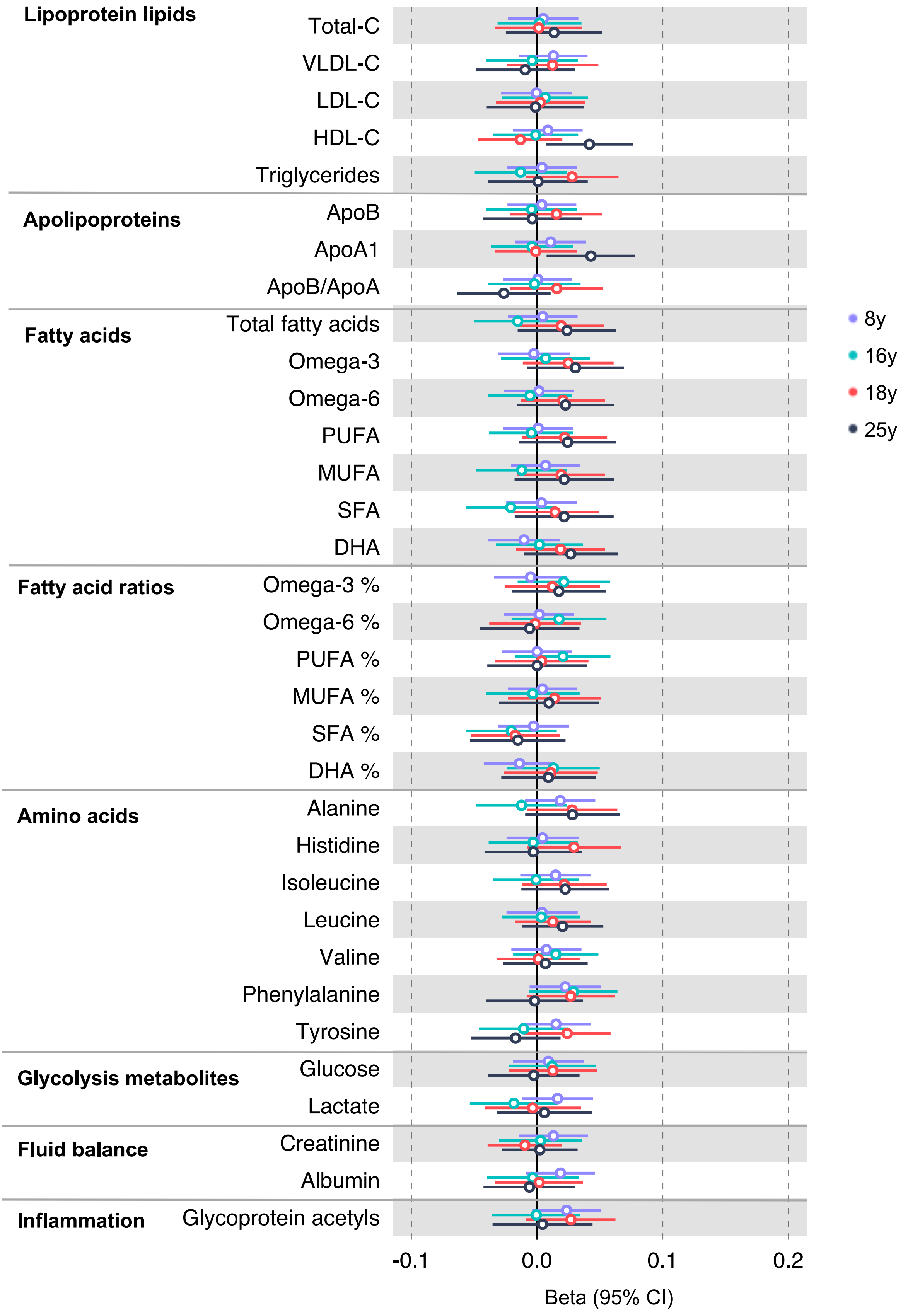

### Figure 2-figure supplement 4

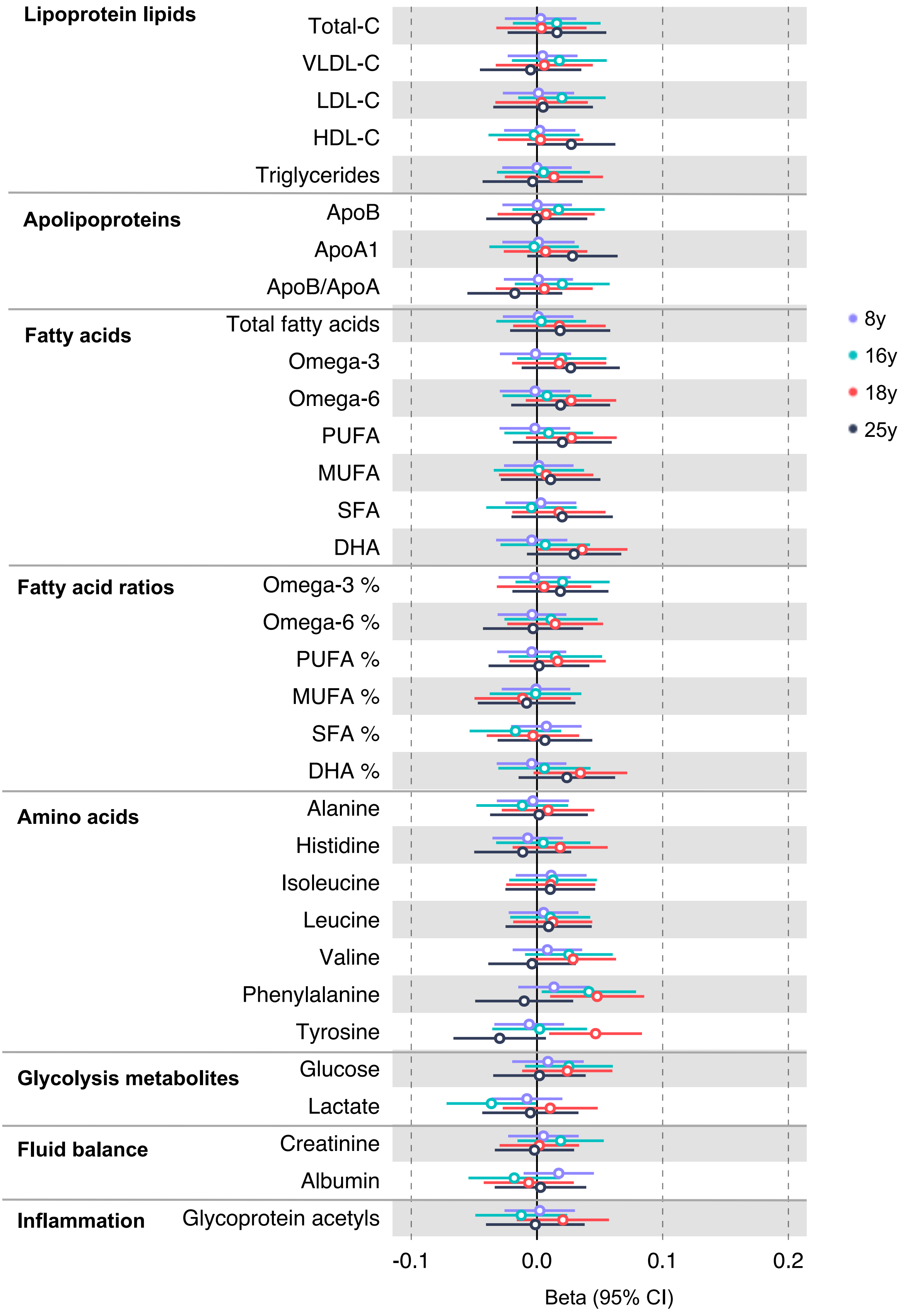

### Figure 2-figure supplement 5

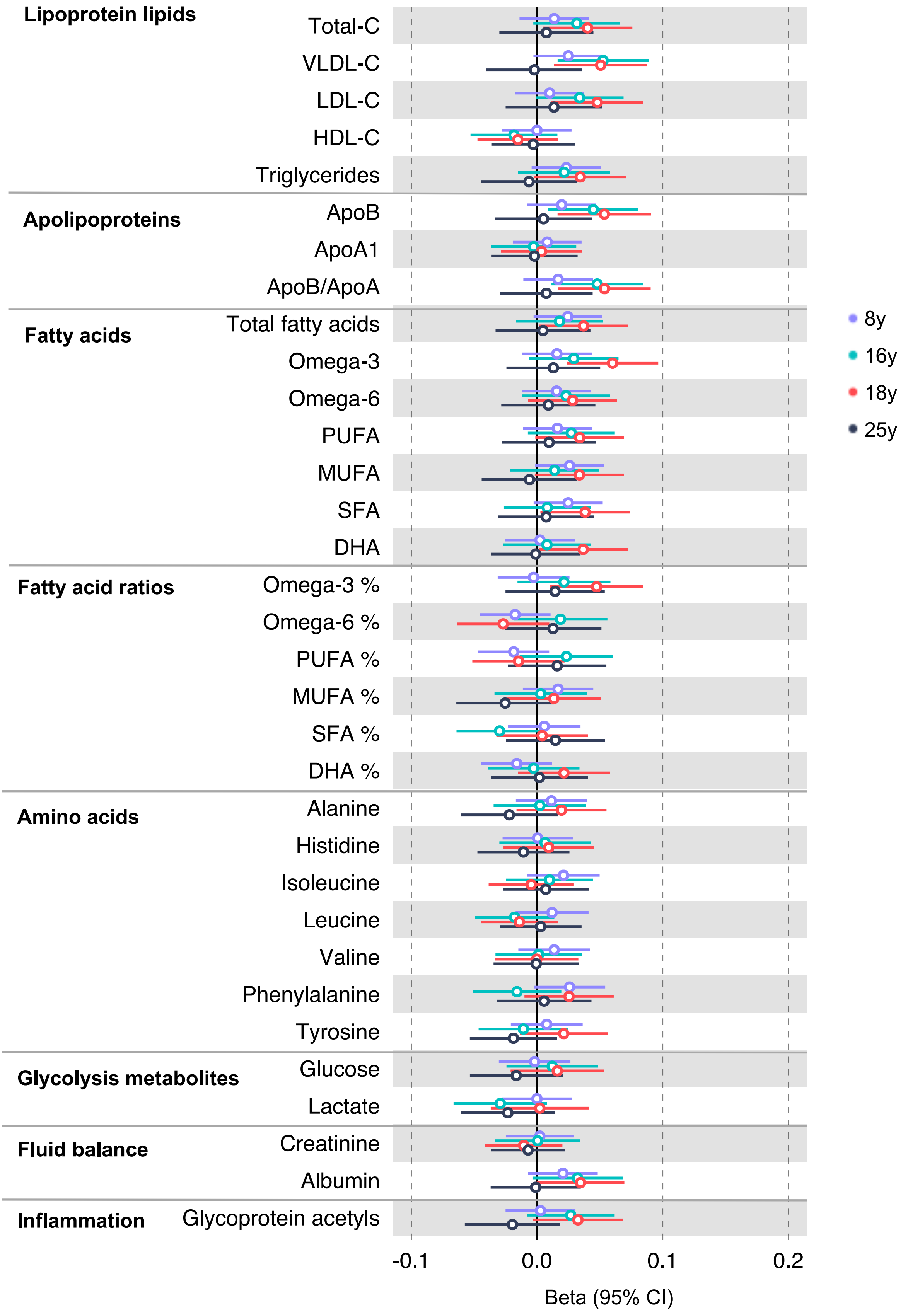

### Figure 2-figure supplement 6

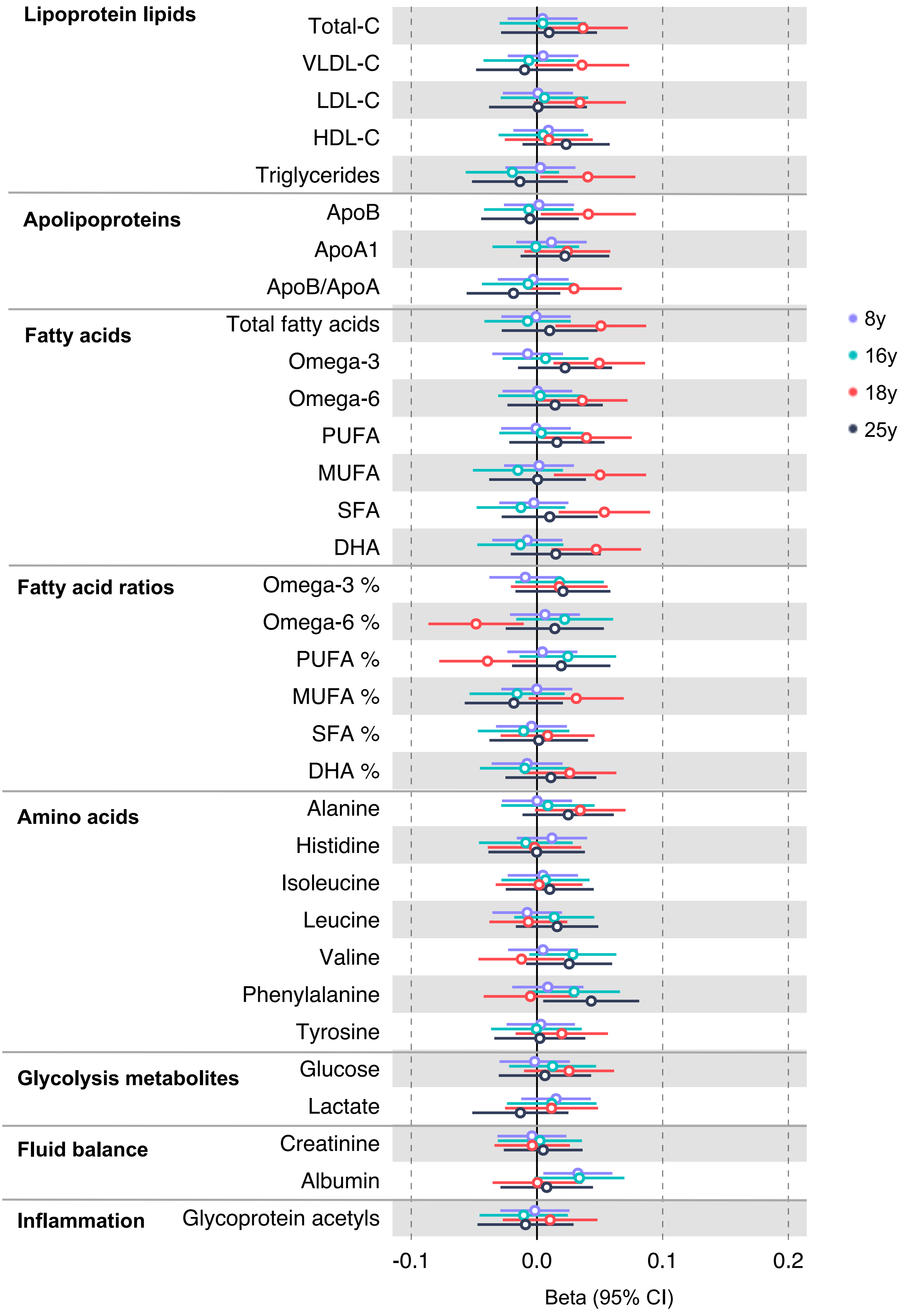

### Figure 3-figure supplement 1

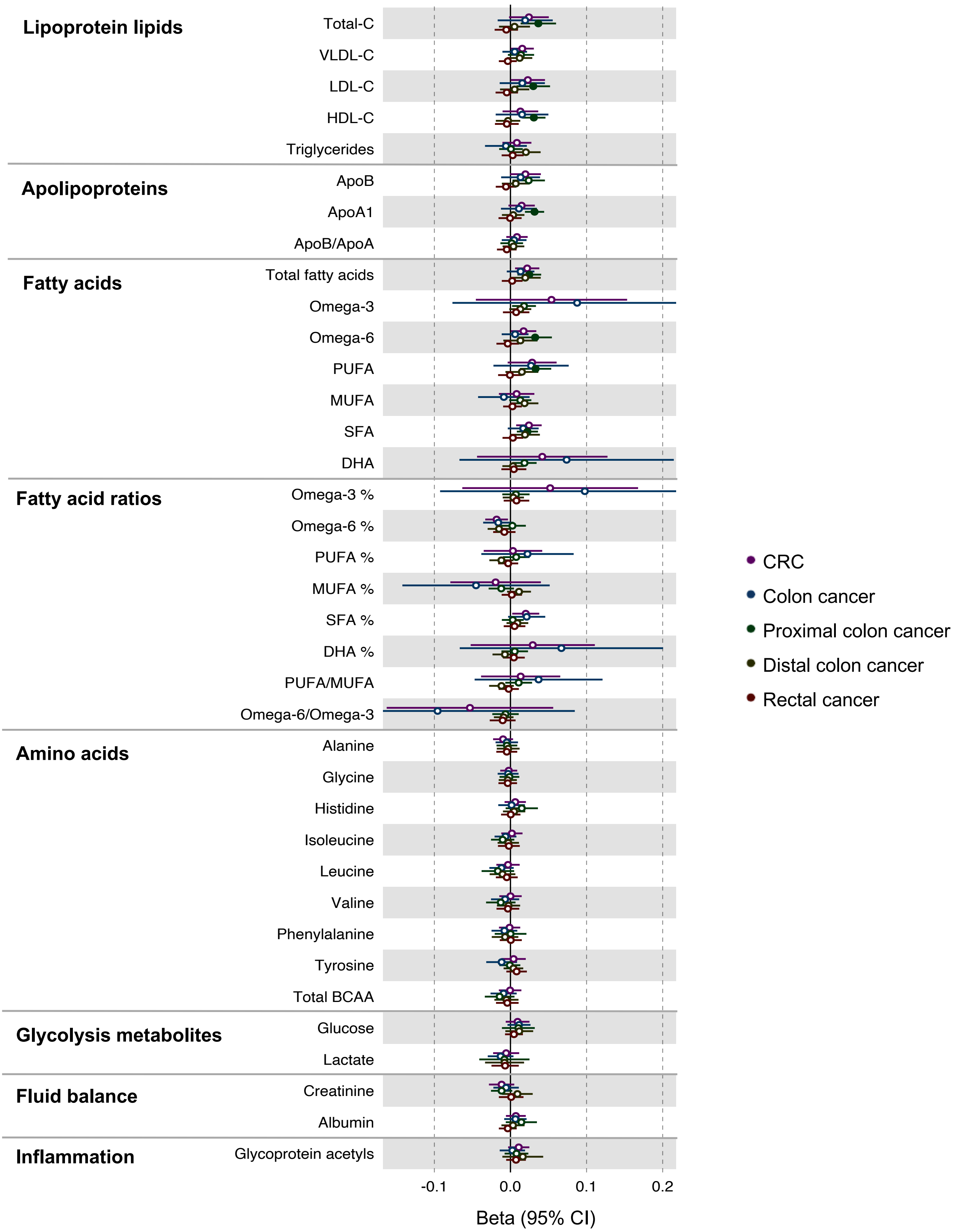

### Figure 3-figure supplement 2

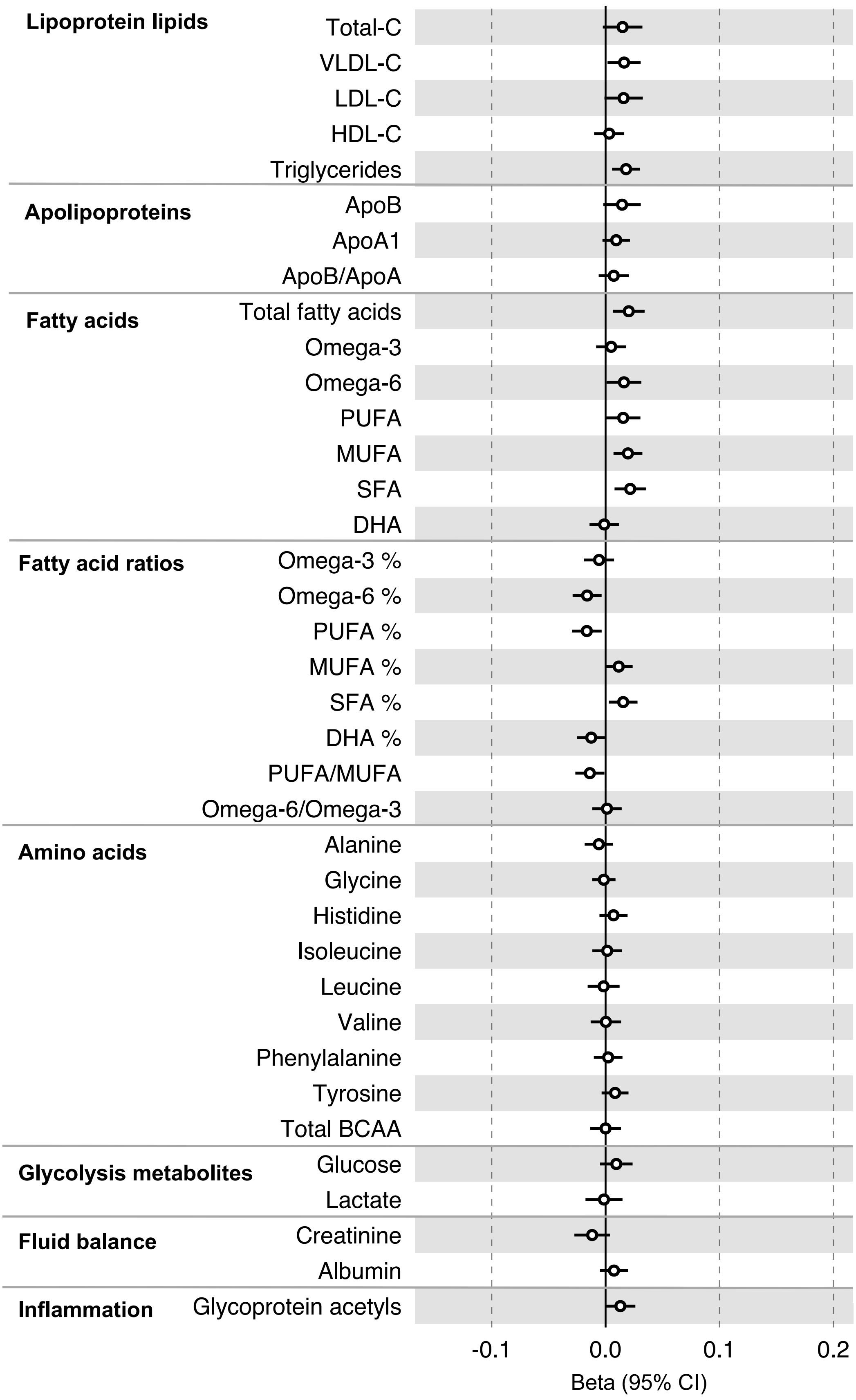

### Figure 3-figure supplement 3

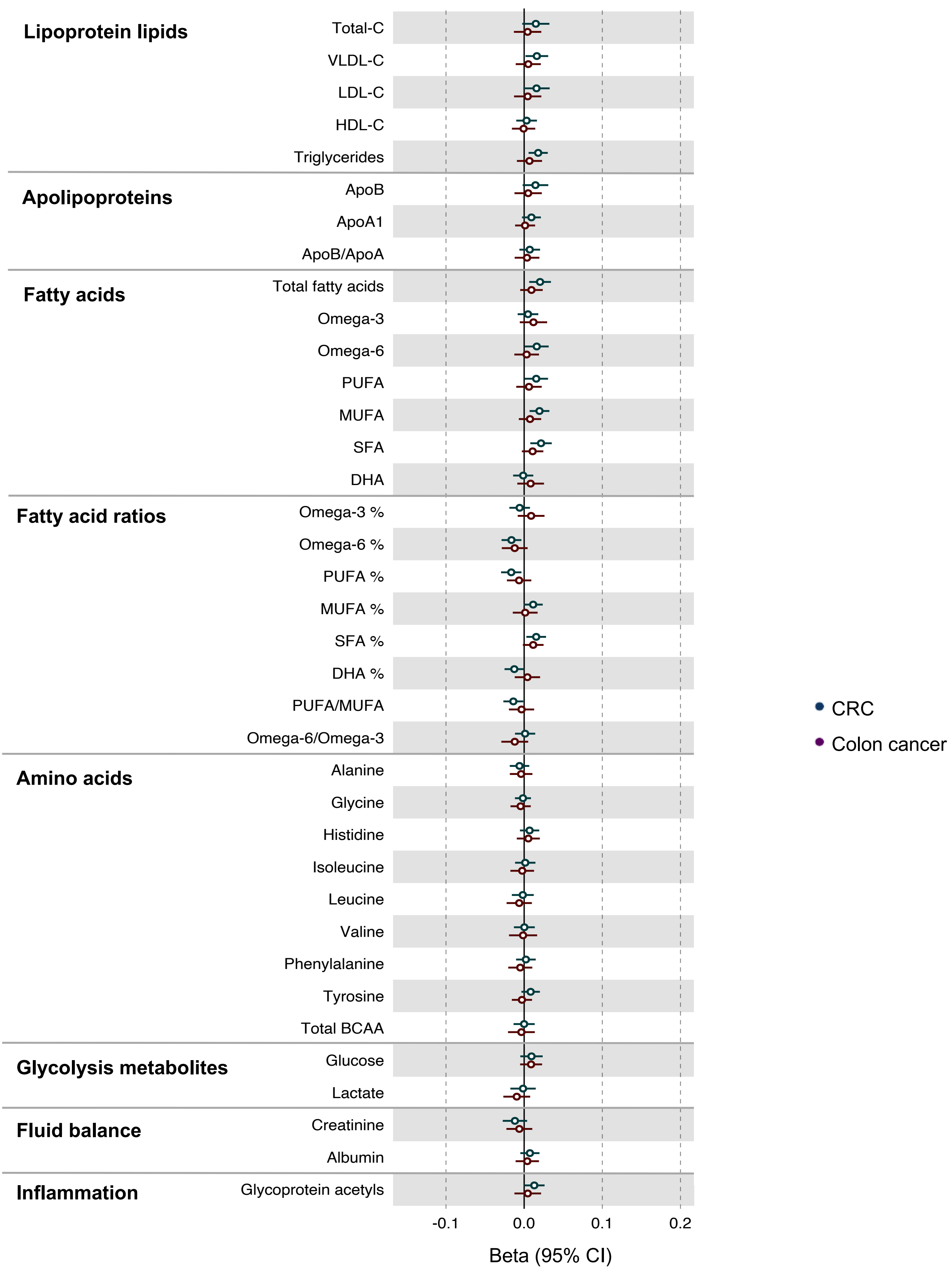

### Figure 4-figure supplement 1

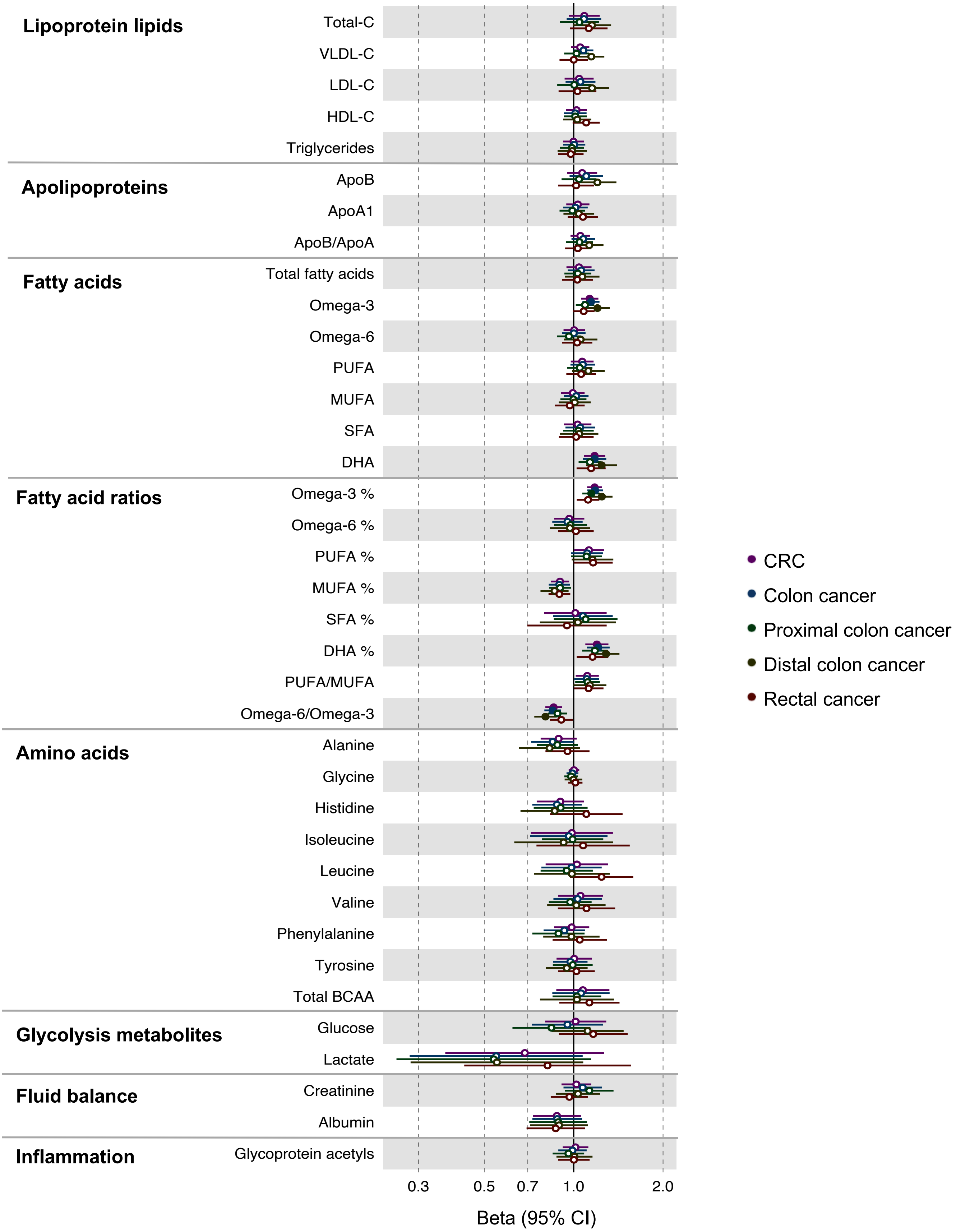

### Figure 4-figure supplement 2

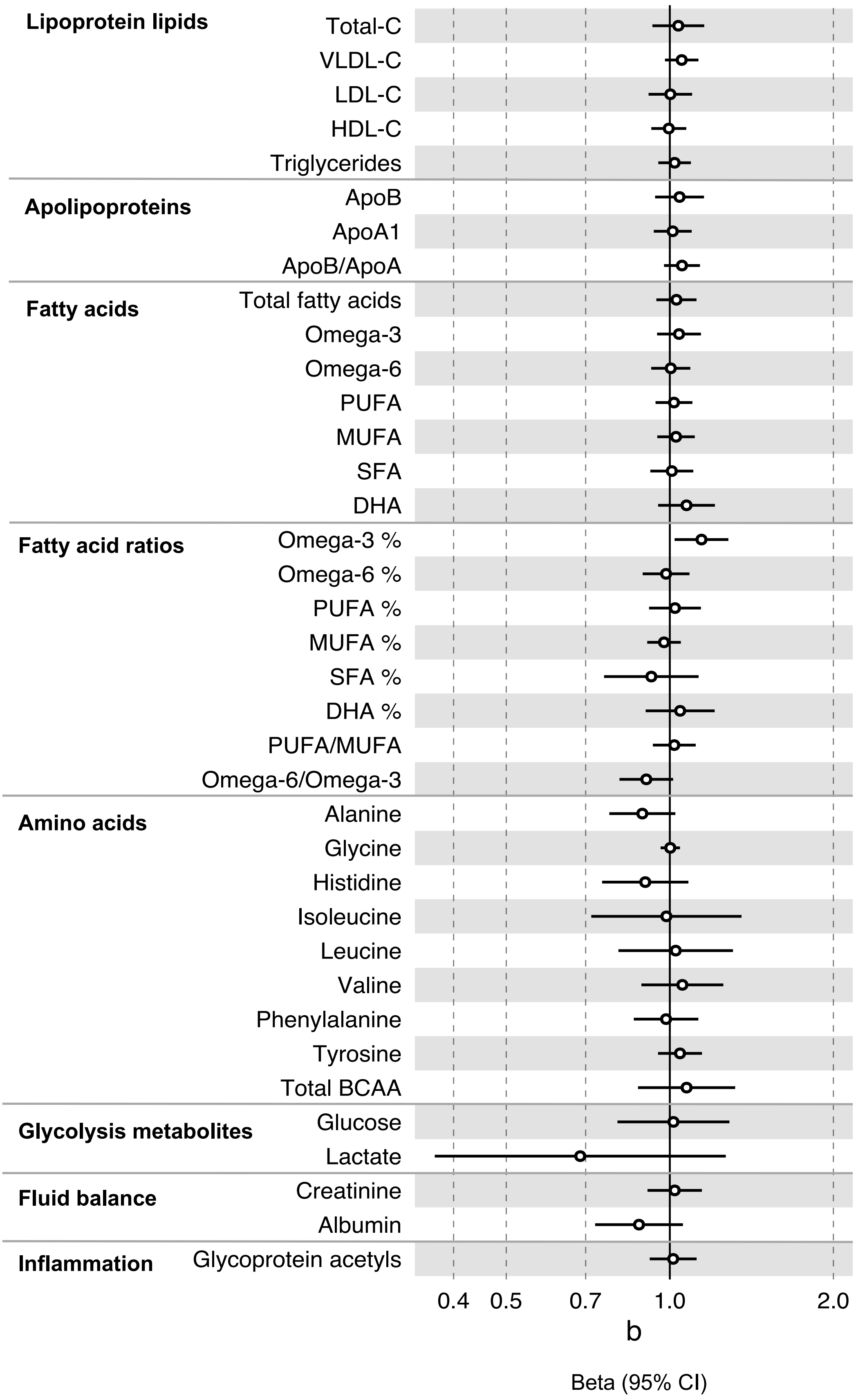

### Figure 4-figure supplement 3

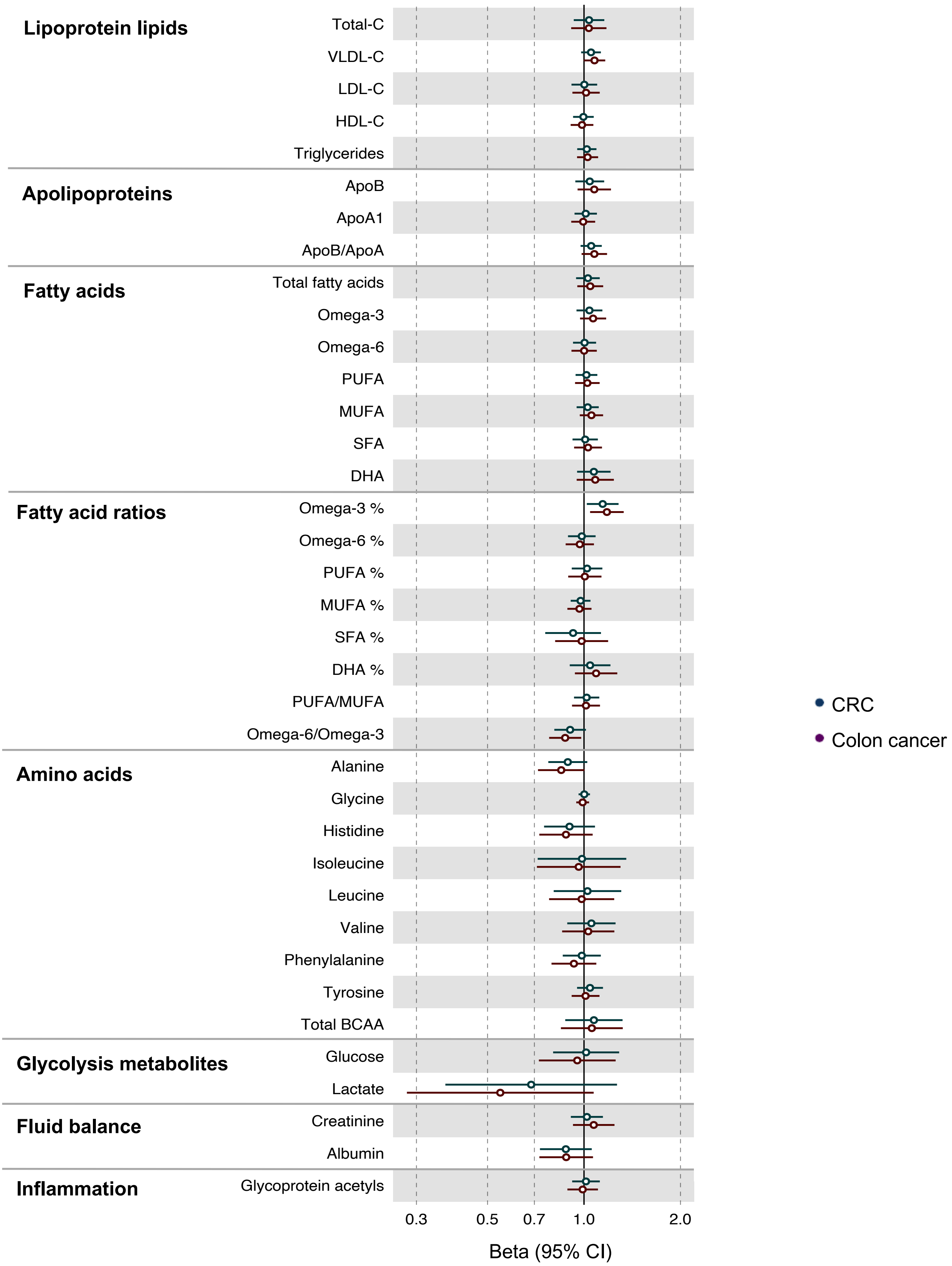
